# VENTILATORY SUPPORT IN SARS-VOC-2 DURING INTENSIVE THERAPY

**DOI:** 10.1101/2020.05.14.20098608

**Authors:** Pereira-Rodríguez Javier Eliecer, Quintero-Gomez Juan Camilo, Lopez-Florez Otilio, Waiss-Skvirsky Sandra Sharon, Velásquez-Badillo Ximena

**Author notes:** The authors declare that they have no conflict of interest. Correspondence to: Javier E. Pereira Rodriguez.

## Abstract

**Introduction:** The SARS-CoV-2 disease outbreak has now become a pandemic. Critical patients with COVID-19 require basic and advanced respiratory support. Therefore, the objective was to describe the ventilatory support strategies in SARS-CoV-2 during intensive therapy.

**Materials and methods:** A systematic review of observational studies of the available scientific literature was performed in accordance with the recommendations of the Cochrane collaboration and the criteria of the PRISMA Declaration.

**Results:** Fifteen observational studies were included that gave a study population of 4,081 patients. Mechanical ventilation is the main respiratory support treatment for critically ill patients, which should be administered as soon as normal oxygenation cannot be maintained, and despite the fact that there is no current consensus on the parameters of mechanical ventilation, the evidence collected suggests the use of Fio2 on average 50%, PEEP of 14 cmH2O, lung compliance of 29-37 ml per cm of water, driving pressure between 12-14 cm of water and a plateau pressure of 22-25 cm of water.

**Conclusions:** IL-6 is shown as a possible marker of respiratory failure and a worse prognosis as well as obesity. In addition, the use of prone position, neuromuscular blockade, pulmonary vasodilators, ECMO, and mechanical ventilation based on the clinical conditions and needs of the patient with COVID-19 are strategies that could benefit patients entering intensive therapy for SARS-CoV-2.

## RESUMEN

**Introducción:** El brote de enfermedad por SARS-CoV-2 actualmente se ha convertido en una pandemia. Pacientes críticos con COVID-19 requieren soporte respiratorio básico y avanzado. Por lo tanto, el objetivo fue describir las estrategias de soporte ventilatorio en SARS-CoV-2 durante terapia intensiva.

**Materiales y métodos:** Se realizó una revisión sistemática de estudios observacionales de la literatura científica disponible de acuerdo con las recomendaciones de la colaboración Cochrane y los criterios de la Declaración PRISMA.

**Resultados:** Se incluyeron 15 estudios observacionales que otorgaron una población de estudio de 4.081 pacientes. La ventilación mecánica es el principal tratamiento de apoyo respiratorio para pacientes críticos, que debe administrarse tan pronto como no se pueda mantener la oxigenación normal y a pesar de que no existe un consenso actual sobre los parámetros de ventilación mecánica, la evidencia recolectada sugiere el uso de Fio2 en promedio del 50%, PEEP de 14 cmH2O, compliance pulmonar de 29-37 ml por cm de agua, driving pressure entre 12-14 cm de agua y una presión plateau de 22-25 cm de agua.

**Conclusiones:** La IL-6 se muestra como un posible marcador de falla respiratoria y peor pronostico al igual, que la obesidad. Además, el uso de posición prona, bloqueo neuromuscular, vasodilatadores pulmonares, ECMO y una ventilación mecanica basada en las condiciones clínicas y necesidades del paciente con COVID-19 son estrategias que podrían beneficar a los pacientes que ingresan a terapia intensiva por SARS-CoV-2.

## INTRODUCCIÓN

SARS-CoV-2 is a coronavirus infection disease recently discovered in humans, which comes from the Coronavirus family and is affecting the world population, becoming a matter of public calamity.

The outbreak of Coronavirus disease was first reported in Wuhan, China on December 31, 2019, and has now become a pandemic, affecting the world population. According to the last report of the World Health Organization (WHO)^1^ for March 30, 2020. 215 territories had reported confirmed cases of COVID-19), 1’655.378 in the Region of the Americas (98.723 deaths), 1‘707.946 in the European Region (155.552 deaths), 255.728 in the Eastern Mediterranean Region (8.878 deaths), 159.662 cases in the Eastern Pacific Region (6.470 deaths), 95.314 in the Southeast Asia Region (3.356 deaths), 42.626 in the African Region (1.369 deaths).

According to the European Center for Disease Prevention and Control, as of May 10, 2020, there were almost 4 million (3’986.119) cases of COVID-19 and 278.814 deaths worldwide. Some of the countries with the highest number of deaths for the same date were: United States (78.794), United Kingdom (31.587), Italy (30.395), Spain (26.478), France (26.310), Brazil (10.627), among others. Likewise, an ECDPC analysis with 17 countries in Europe determined that the most commonly reported clinical symptoms were fever (47%), dry or productive cough (25%), sore throat (16%), general weakness (6%) and pain (5%); hospitalization occurred in 30% (13.122 of 43.438) of the cases reported in 17 countries, and the cases requiring ICU or respiratory support represented 2.179 of 49.282 (4%)^2^.

Understanding that there is still no clarity on the treatment for patients with Coronavirus, it has been described based on the pathophysiology of COVID-19 requiring advanced life support needs, ranging from oxygen supplementation through non-invasive ventilation, to invasive mechanical ventilation and vasopressor support; some secondary complications described in confirmed cases include cardiomyopathy, pulmonary embolism, and sudden death^3,4^.

Based on the available data, it is defined that the viral infection is capable of producing an excessive immune reaction in the host, generating in turn what is called “cytokine storm”, an effect of extensive tissue damage in cascade; Interleukin 6 (IL-6) is the main proinflammatory protein produced by leukocytes in cases of COVID-19, and which act on a large number of cells and tissues. In this way, and unlike other types of Coronavirus described above, SARS-CoV-2 has shown greater systemic involvement than others, mainly of respiratory origin associated with pneumonia^5,6^.

In one of the first reports on the disease, chest computed tomography (CT) was performed on patients with COVID-19, where pneumonia with abnormal findings was evident in all cases. About a third of them (13.32%) required intensive care unit (ICU) care, and there were 6 (15%) fatal cases. From this, the needs for assistance and support in the ICUs of all the countries of the world has increased. Likewise, each time the cases of acute respiratory distress syndrome (ARDS) and SARS-CoV-2-associated pneumonia continue to grow, which has forced the assistance of mechanical ventilation in these cases^7,8,9^.

A report by the Intensive Care National Audit and Research Center (ICNARC)^10^ in the United Kingdom described the results of an observational cohort study with 165 patients in critical care units. Of the 165 patients, 79 (47.9%) died and 86 (52.1%) were discharged alive from the ICU. On the other hand, 58.8% of the patients required advanced respiratory support, 50.3% basic respiratory support, 20.6% advanced cardiovascular support, 84.2% basic cardiovascular support, 17% renal support and 2.4 % required neurological support.

The percentage of patients with SARS-CoV-2 who require advanced ventilatory support, such as mechanical ventilation (MV), is high. However, there are currently no specific protocols for the management of MV in these patients. On the other hand, the need for updating based on scientific evidence on the assistance and management of mechanical ventilation in patients with SARS-CoV-2 has increased. Thus, this study aims to describe the ventilatory support strategies in SARS-CoV-2 during intensive therapy.

## MATERIALS AND METHODS

### Design

A systematic review was performed with a descriptive analysis of retrospective chronology of clinical trials, case reports, systematic reviews and meta-analyzes, available and published in indexed databases. Research that included experimental studies with human beings had informed consent under the ethical considerations of Helsinki^11^, for the regulation of experimental studies in living beings.

### Search strategy

The search for scientific documents was developed under the considerations and criteria of the PRISMA Flow Chart^12^ (Transparent Reporting Items for Systematic Reviews and Meta-Analyzes) for the identification, screening, eligibility and inclusion of studies in systematic reviews. Different databases were reviewed for the identification of prospective documents based on the criteria of the PRISMA Diagram for text search. These search criteria are defined as: records identified through the database search, additional records identified through other sources, records after duplicates have been removed, selected records, full-text articles evaluated for eligibility, and finally studies included in qualitative synthesis. The databases identified for searching scientific studies included: PubMed and PubMed Central.

### Boolean descriptors and operators

Descriptors associated with the variables described in the study title were used: mechanical ventilation & COVID-19; and the boolean operators: AND & OR. Thus, the search strategy was as follows: (“respiration, artificial” [MeSH Terms] OR (“respiration” AND “artificial”) OR “artificial respiration” OR (“mechanical” AND “ventilation”) OR “mechanical ventilation” [) AND (“COVID-19” OR “COVID-2019” OR “severe acute respiratory syndrome coronavirus 2” OR “severe acute respiratory syndrome coronavirus 2” OR “2019-nCoV” OR “SARS-CoV-2” OR “2019nCoV “OR (((“ Wuhan “AND (“ coronavirus “[MeSH Terms] OR” coronavirus “)) AND (2019/12 OR 2020))).

### Selection of studies

The selection of articles was made by the different authors of the study. At first, one author (JCQG) identified prospective studies, later a second author (OL-P) carried out the screening or screening process, a third researcher (SSW-S) applied eligibility criteria, and finally full text analysis and included by previous collaborators. To reduce study selection and inclusion bias, each of the research collaborators independently reviewed the studies chosen to analyze the full text using the PRISMA Checklist^13^, to finally agree on the studies to include in the sample (n). On the other hand, a fourth researcher.

### Selection criteria

Eligibility criteria were defined in the PRISMA Checklist, according to the design characteristics of the included studies and characteristics of the reported population. Among the general criteria applied to all the studies, the place of publication, year of publication, type of literature, language of publication, study design, methodology, nature and characteristics of the population were taken into account.

From the above, studies published in indexed databases, documents available to date, scientific article-type texts, experimental and descriptive design research on the application and management of mechanical ventilation in humans with coronavirus acquired in the current pandemic. Some data such as sex, age, anthropometric characteristics, ethnicity and comorbidities were not filtered; one author (JEPR) verified that the ethical recommendations were complied with in each of the included investigations; all the collaborators verified the application of inclusion criteria.

### Data collection and extraction

The data were extracted into a selection matrix for the identified, screened, chosen and finally included studies, designed by the authors through “Excel” spreadsheets. Data from the studies chosen in the systematic review were collected on information analysis sheets. The information on the studies included in the review was described using characterization tables on text sheets.

### Type of participants

Subjects of both genders, from any region of the world with confirmed diagnosis for COVID-19, who received supplemental oxygen from invasive or non-invasive mechanical ventilation were included.

## RESULTS

### Selection of studies and characteristics

After the initial search, 1,128 titles were identified regarding the use of mechanical ventilation in patients with COVID-19. A total of 104 documents were selected that described cases of patients who required mechanical ventilation due to the severe hypoxemia generated by the coronavirus. Subsequently, 64 studies were chosen for full-text review (study objective, interventions carried out, results obtained and conclusions).

Finally, 15 observational studies were included^15-29^ (**figure 1**) after the full text review, on mechanical ventilation in COVID-19 that were published between February 9, 2020 and April 15, 2020; 3 clinical cohort studies^20,26,27^ and 12 retrospective clinical studies^15-19,21-25,28,29^. In this way, the sample size (n) of each included study, the main diagnosis, the information collection methodology, the time, results and conclusions of each study included in the systematic review were determined (**table 1**).

**Fig. 1.**
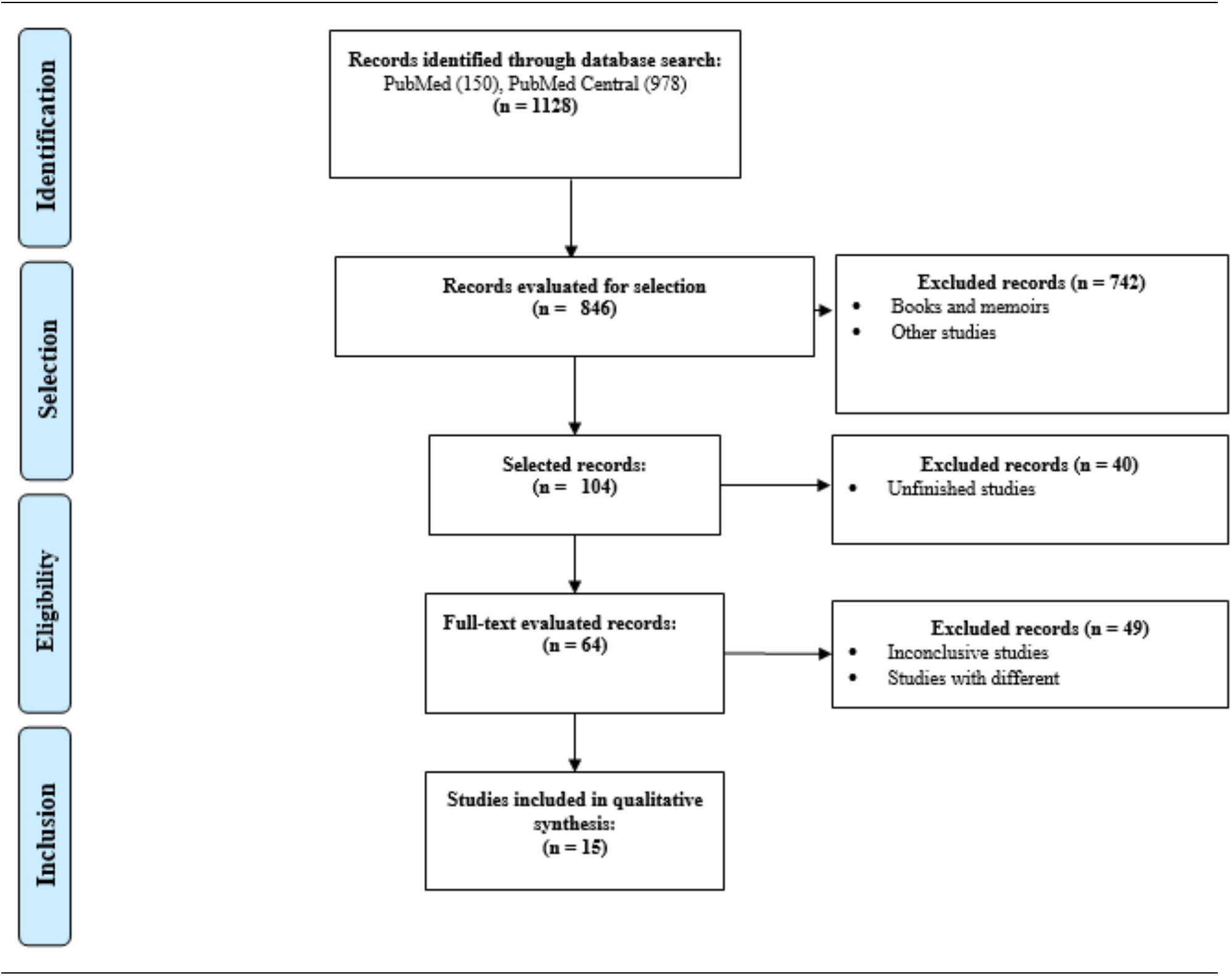
Illustration of the PRISMA Flowchart for the Identification, selection, eligibility and inclusion of studies in systematic reviews.

**Table 2.**
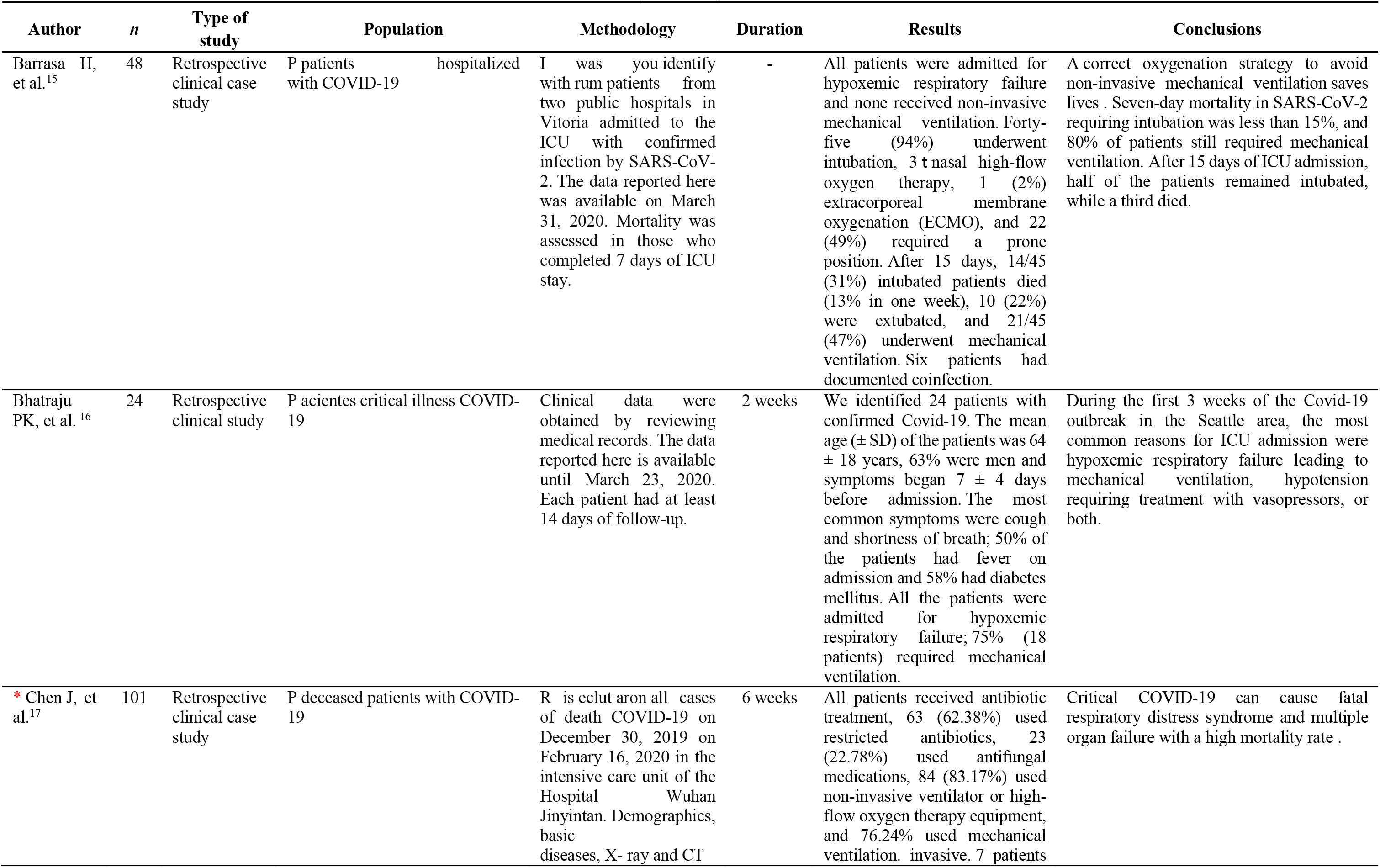
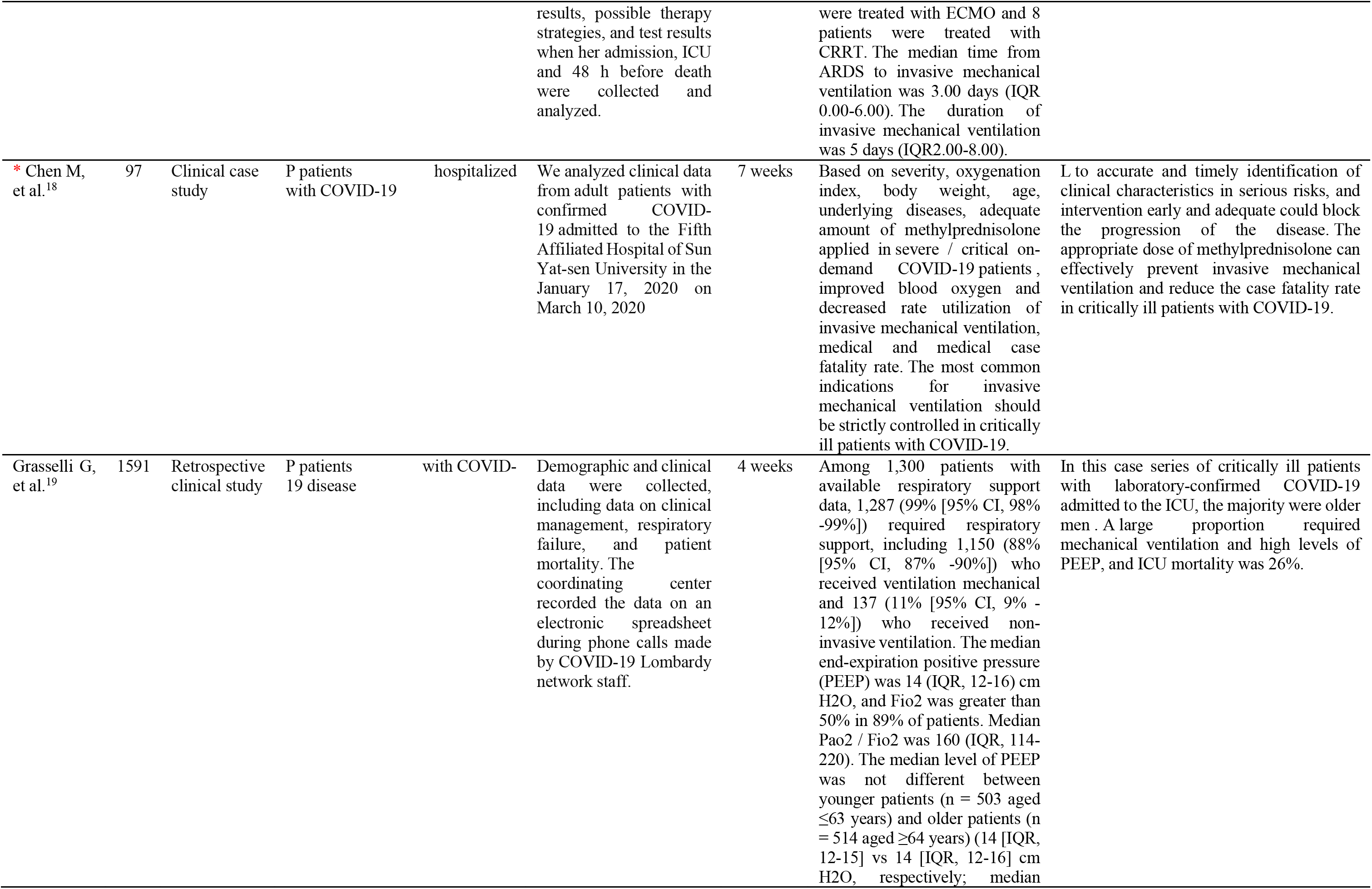
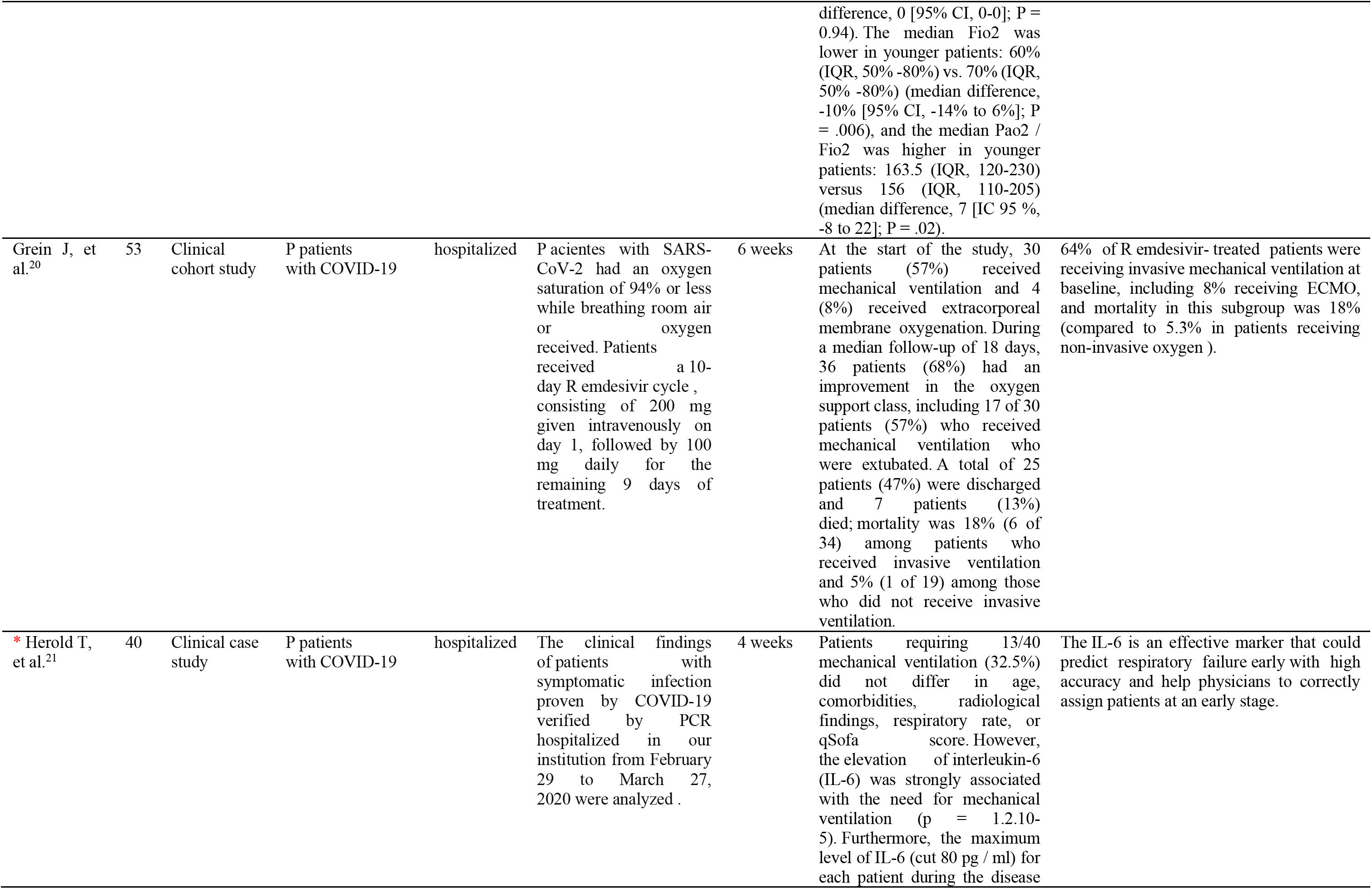
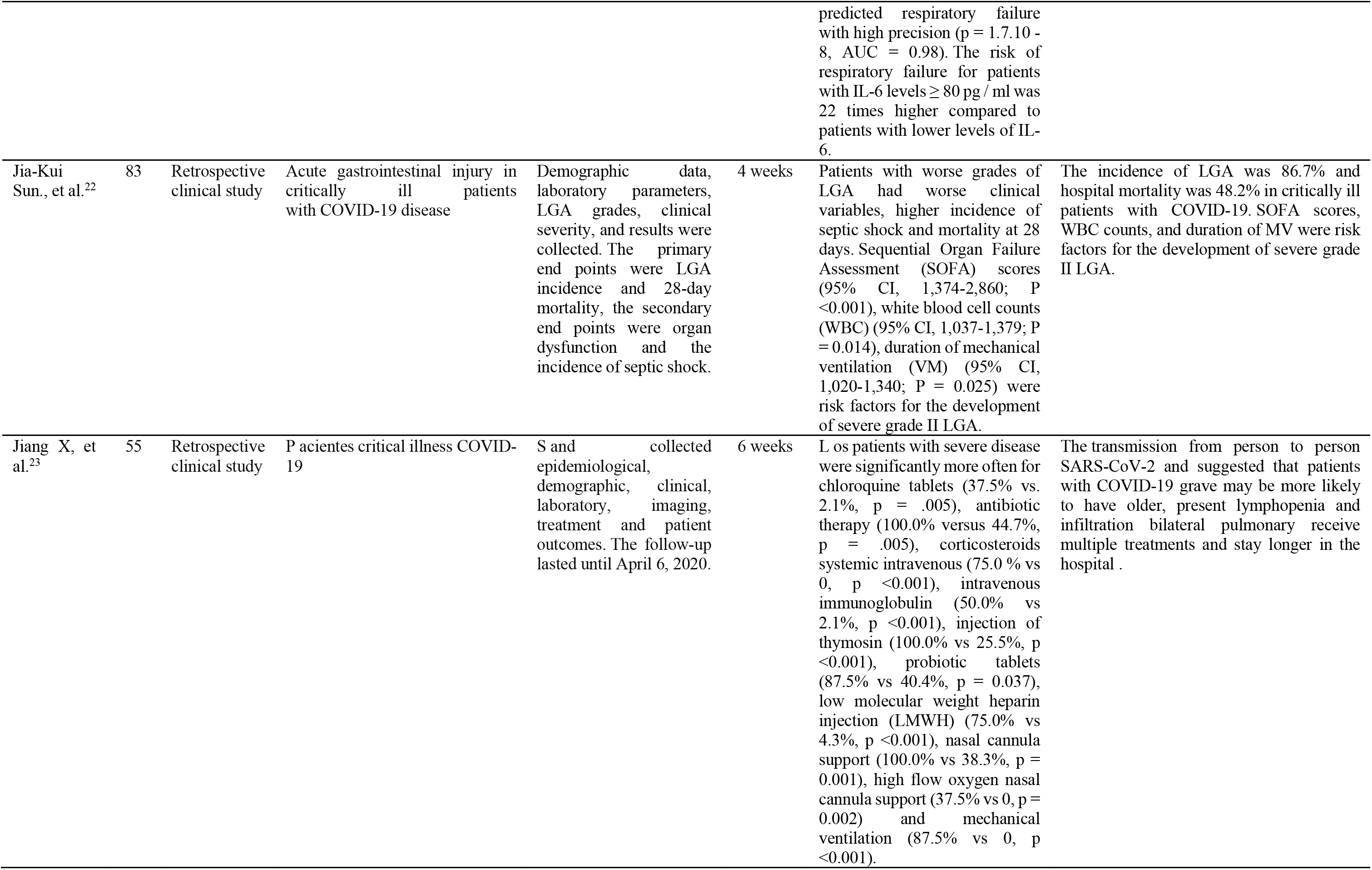
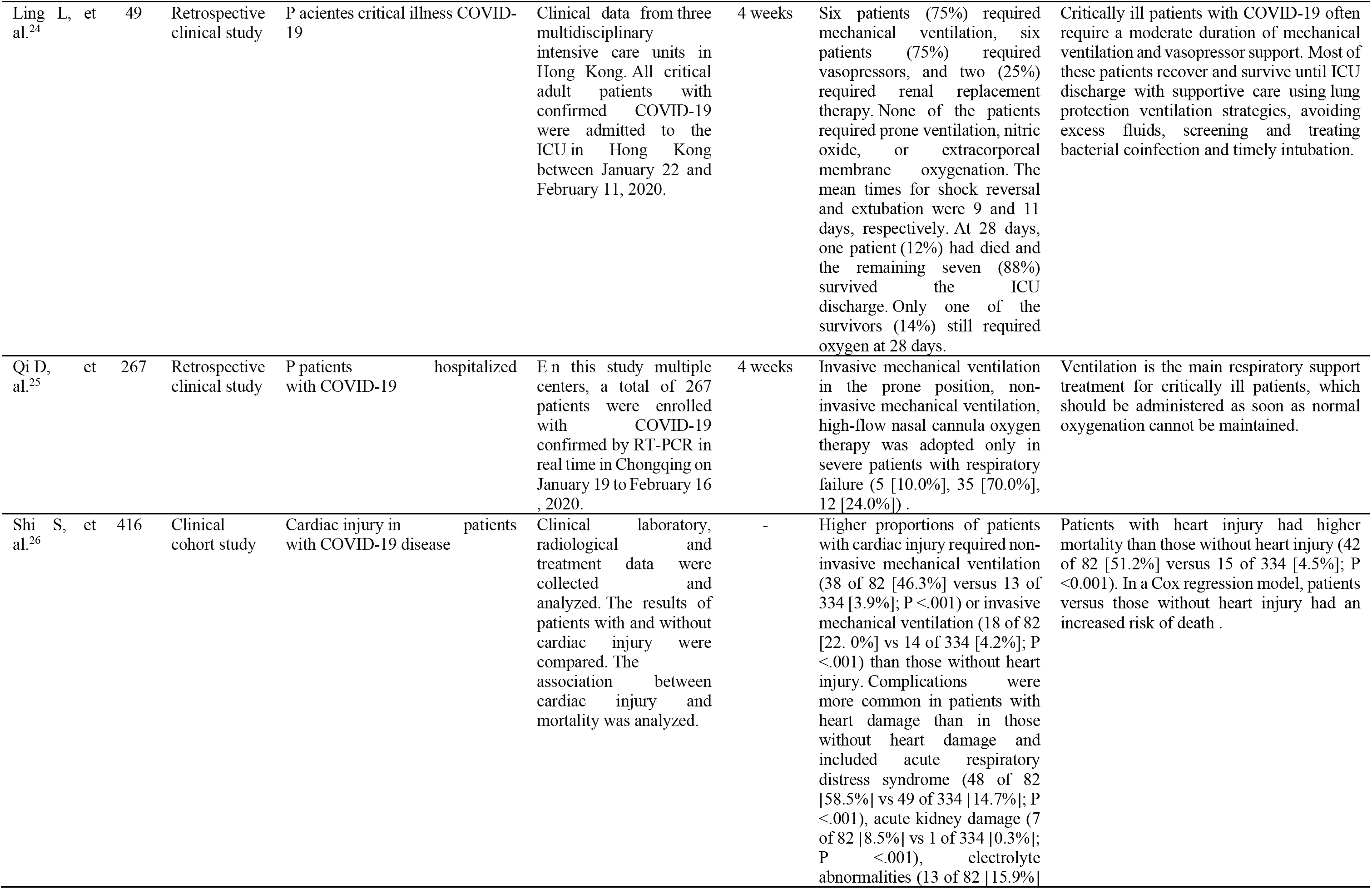
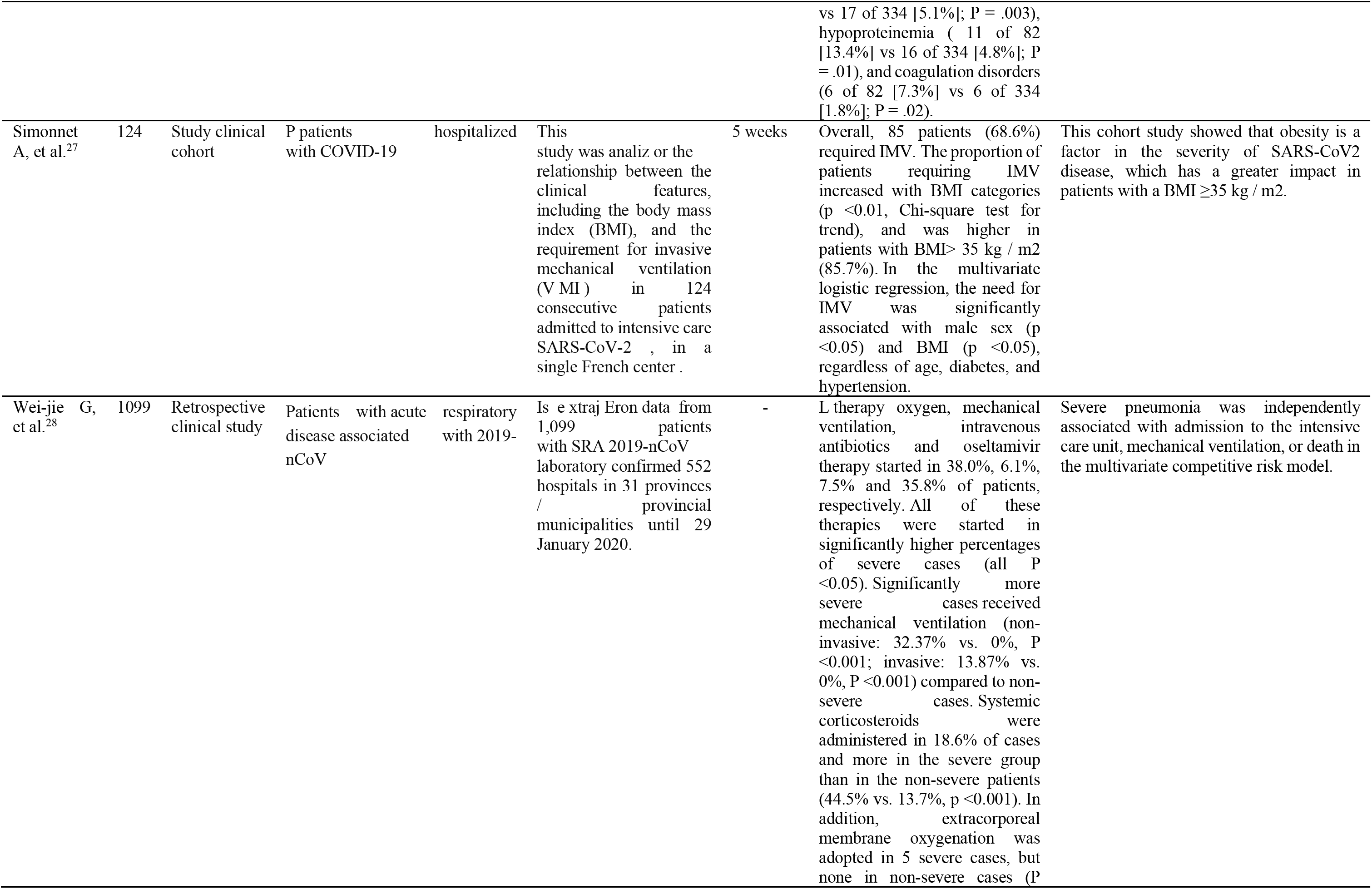
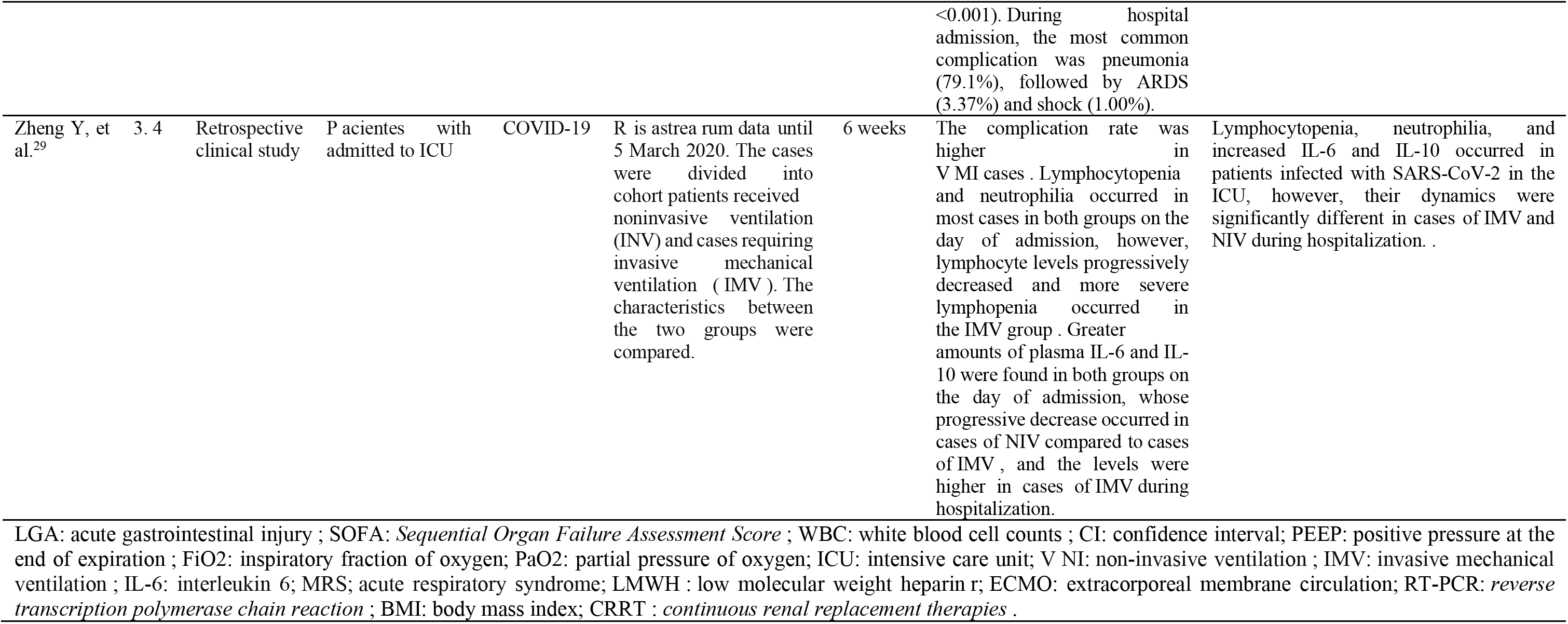
Characteristics of the interventions of the included studies (*n* = 1 5)

### Mechanical ventilation in patients with COVID-19

Mechanical ventilation is the main respiratory support treatment for critically ill patients, which should be administered as soon as normal oxygenation cannot be maintained^25^. The use of mechanical ventilation in critically ill COVID-19 patients remains a strategy for improving ventilation and mitigating the impact on acute respiratory distress syndrome developed by many patients with coronavirus. Currently, there is little evidence on the needs for MV in patients with coronavirus and the prognosis for improvement. Critically ill patients with COVID-19 often require a moderate duration of mechanical ventilation and vasopressor support^24^.

### Strategies

Invasive mechanical ventilation in the prone position^15,16,26^, non-invasive mechanical ventilation^15-17,19,21,23,24,26-29^, high flow nasal cannula oxygen therapy^15,23,26^ and extracorporeal membrane oxygenation (ECMO)^15,17,20,29^, are some strategies in critically ill patients with SARS-CoV-2 who develop respiratory failure; as well as the use of neuromuscular blockade, inhaled pulmonary vasodilators and vasopressors^16^.

### Mortality

On the other hand, a high percentage of patients with SARS-CoV-2 who enter the ICU require prolonged mechanical ventilation^15,16,17^ and a higher percentage requires invasive mechanical ventilation^15-17,19,20,23,24,25-28^. Mortality in patients with invasive mechanical ventilation (~18% to 33,3%) seems to be greater compared to non-invasive mechanical ventilation (~5% a 10%)^15,17^ and even behaves up to 26%^19^.

### Parameters

Although there is no current consensus on the parameters of mechanical ventilation in patients with COVID-19, most studies suggest the volume-controlled / assisted ventilatory mode, the use of elevated Fio2 from day 1 of mechanical ventilation, among 60% and 90%^16,19^, although the mean in the different studies was Fio2 50%^19^, a *pulmonary compliance* to 29-37 ml for cm to water^16^, positive pressure at the end of expiration (PEEP) of 14 (IQR, 12-16) cm H2O, plateau pressure of less than 30ml H2o, a *driving pressure* (the difference between the plateau pressure to the PEEP) of 12-14 cm of water^16,19^ and a pressure *plateau* of 22-25 cm of water^1616,19^ are the guidelines recommended by the evidence collected.

### Comorbidities and injuries

Patients with cardiac injury and COVID-19 require more mechanical ventilation support than those without cardiac injury^26^; 46.3% non-invasive and 22% invasive mechanical ventilation^26^. On the other hand, higher values of body mass index (IMC> 35 kg / m2) are associated with higher invasive mechanical ventilation requirements^27^. Similarly, higher levels of systemic blood pressure are associated with higher demand for MV and higher mortality^19,22,27^.

Furthermore, the elevation of interleukin-6 (IL-6) is strongly associated with the need for mechanical ventilation. Furthermore, the maximum level of IL-6 (cut 80 pg / ml) for each patient during the disease predicts respiratory failure with high precision. The risk of respiratory failure for patients with IL-6 levels ≥ 80 pg / ml is 22 times higher compared to patients whose levels of IL-6 are lower. Also, the incidence of acute gastrointestinal injury (LGA) in critically ill patients with COVID-19 undergoing prolonged MV is frequent (86.7%)^21^.

Lymphocytopenia and neutrophilia occur in most cases, however, lymphocyte levels progressively decrease and lymphopenia occurs, which is more severe in patients with IMV^29^. Likewise, the rate of patients undergoing continuous renal replacement therapy for multiple organ failure and kidney injury in patients with SARS-CoV-2 is high^17,24,26^. Furthermore, critical COVID-19 can cause fatal respiratory distress syndrome and multiple organ failure with a high mortality rate17 and very frequently pneumonia (79.1%), followed by ARDS (3.3%) and shock (1.0%)^28^.

### Other strategies

The use of methylprednisolone applied in critically ill COVID-19 patients appears to improve blood oxygen, reduce the rate of use of IMV, and the mortality rate^18^.

## DISCUSSION

This systematic review highlights the needs for basic and advanced respiratory support in patients with SARS-CoV-2. The respiratory care in COVID-19 described by Meza, C. et al. (2020)^30^ seeks to generate lung protection strategies by decreasing tidal volumes, plateau pressure, respiratory rates, driving pressure values and prone ventilation, as well as implementing high PEEP values, which have shown improvement in hypoxemia and survival in patients with acute respiratory distress syndrome, the same characteristics described in this article based on the needs for protective mechanical ventilation in patients with coronavirus and with a prognosis for improvement.

On the other hand, other statements^31^ mention that mechanical ventilation is capable of producing and aggravating lung damage by the administration of supplemental oxygen in inadequate amounts, highlighting that the effects of hyperoxia in the lungs can lead to the formation of alveolar hyaline membrane, edema, hyperplasia, proliferation of type II pneumocytes, destruction of type I pneumocytes, interstitial fibrosis and pulmonary vascular remodeling, data that resemble those proposed in the present investigation, associating that the inadequate dosage of oxygen according to the patient’s clinical conditions would result counterproductive in its evolution.

Likewise, Vidal, F. & Calderón, V. (2012)^32^ affirm that mechanical ventilation in acute respiratory distress syndrome not only involves respiratory disorders but also supposes an elevation of alveolar and transpulmonary pressure, conditioning a significant alteration and overload. for the function of the right ventricle that can fail giving rise to the clinical picture of acute pulmonary cor, it is for this reason that the importance of continuous monitoring of pulmonary and hemodynamic mechanics is highlighted when considering ventilatory strategies; likewise, as mentioned by Gattinoni L. et al. (2020)^33^ referring to the phenotypes of SARS-CoV-2 associated pneumonia. In this way, it is reported in a study^34^ that the prone position has a great impact on cardiopulmonary physiology, being a useful and accessible maneuver for most intensive care units, and the findings are also mentioned in the present investigation. relevant of this technique on terms of survival in patients with relatively severe ARDS (PaO2 / FiO2 ≤ 150mmHg) highlighting that it is necessary to reevaluate the levels of PEEP once the maneuver has been performed, adjusting to the particularities of each clinical situation.

In another study^35^, they mention that the use of muscle relaxants in the hypoxemic patient seeks to improve patient-ventilator synchrony, resulting in conflict with the development of myopathy as well as reducing the benefits of spontaneous breathing, as it was also shown that muscle relaxants, in patients with ARDS criteria treated under deep sedation showed an improvement in thoracic compliance and a decrease in O2 consumption.

That said, the use of non-invasive mechanical ventilation is questioned due to the findings found for its use in patients with ARDS as described by Franca, A. et al. (2014)^36^ showing a trend of higher failure and mortality in hypoxemic respiratory failure, also emphasizes that values of respiratory rate >30 rpm in the first hour of NIV are associated with failure in hypoxemic and hypercapnic respiratory failure accompanied by a greater number of infectious complications. Therefore, its use is limited to the different respiratory situations of the patient in which its placement is valued by the respiratory conditions as well as its hemodynamic stability described in the present investigation.

Consequently, Cristancho, W. (2020)^37^ mentions the use of extracorporeal membrane oxygenation (ECMO) as an alternative to respiratory care, stating that its early use did not significantly improve mortality in patients with severe ARDS, however when used as a modality of rescue could help improve survival in patients with acute respiratory syndrome. And what has been said in this research, added to the corroborative evidence that we find from our findings, is to highlight that the actions in intensive care are fundamental for the survival of patients whose consequences have already been mentioned, together with prolonged periods of immobilization and bed rest, among which are those mentioned by the Pan American Health Organization^38^: Impaired lung function; physical deconditioning and muscle weakness; cofusional symptoms and other cognitive deficiencies; dysphagia and difficulties to communicate; mental health disorders and need for psychosocial support. Therefore, the need for rehabilitation professionals who play an important role in ICUs by facilitating early discharge is considered, which is especially important in a context of scarcity of hospital beds. Likewise, to prevent and intervene in sequelae associated with severe COVID-19. For this reason, rehabilitation professionals should be assigned to ICUs, hospital wards, transition facilities and the community. However, it should be noted that the limitations of this study refer to the limited scientific evidence on the use of mechanical ventilation in patients with SARS-CoV-2, however, different authors suggest, like this review, that the Invasive mechanical ventilation is associated with a worse prognosis and a higher percentage of mortality when not used correctly and with academic grounds.

Similarly, the importance of very good early advanced ventilatory support is highlighted, since it can avoid the use of IMV in patients with COVID-19^39-43^.

## CONCLUSIONS

All SARS-CoV-2 patients require respiratory support and a very high rate requires mechanical ventilation. IL-6 is shown as a possible marker of respiratory failure and a worse prognosis, like that of patients with obesity. In addition, the use of prone position, neuromuscular blockade, pulmonary vasodilators, ECMO, and mechanical ventilation based on the clinical conditions and needs of the patient with COVID-19 are strategies that could benefit patients entering intensive therapy for COVID-19. Likewise, it is highlighted that the use of methylprednisolone seems to reduce mortality and hospital stay.

## Data Availability

We have all the information presented in this investigation and we leave it at the disposal of the different authors and that they consider important to be able to replicate or, in effect, use it for their investigations.

